# Assessing the potential impact of COVID-19 in Brazil: Mobility, Morbidity and the burden on the Health Care System

**DOI:** 10.1101/2020.03.19.20039131

**Authors:** Flávio C Coelho, Raquel M Lana, Oswaldo G Cruz, Daniel Villela, Leonardo S Bastos, Ana Pastore y Piontti, Jessica T Davis, Alessandro Vespignani, Claudia T Codeço, Marcelo F C Gomes

## Abstract

**Background:** Brazil detected community transmission of COVID-19 on March 13, 2020. In this study we identify which areas in the country are most vulnerable for COVID-19, both in terms of the risk of arrival of COVID-19 cases and the risk of sustained transmission. The micro-regions with higher social vulnerability are also identified.

**Methods:** Probabilistic models were used to calculate the probability of COVID-19 spread from São Paulo and Rio de Janeiro, according to previous data available on human mobility in Brazil. We also perform a multivariate cluster analysis of socio-economic indices to identify areas with similar social vulnerability.

**Results:** The results consist of a series of maps of effective distance, outbreak probability, hospital capacity and social vulnerability. They show areas in the North and Northeast with high risk of COVID-19 outbreak that are also highly vulnerable.

**Interpretation:** The maps produced are useful for authorities in their efforts to prioritize actions such as resource allocation to mitigate the effects of the pandemic and may help other countries to use a similar approach to predict the virus route in their countries as well.

**Funding:** No funding

## 1. Introduction

As of 21 March 2020, the pandemic of COVID-19 has reached 184 countries with 266,073 confirmed cases and 11,184 deaths, globally [1]. The first imported case of COVID-19 was confirmed in Brazil on February 26, 2020, in the city of São Paulo [2], only two months after the alert on COVID-19 went off in China. All twenty seven federative units currently have suspect cases of COVID-19 infection, while 16 states and the Federal District have already confirmed a total of 428 cases (11,278 under investigation) [3].

São Paulo and Rio de Janeiro states identified virus community transmission on March 13, 2020 [4, 5, 6]. Just four days later, 240 cases were confirmed in São Paulo, 4 deaths were registered, and 5,334 individuals remain under investigation [3]. In Rio de Janeiro, 45 have been confirmed and 1,254 individuals are under investigation, according to the Ministry of Health [3]. The corresponding capitals – which are also called São Paulo and Rio de Janeiro – are the most populous metropolitan areas of Brazil, with millions of people living in precarious conditions. These two states are also the country’s main local and international hubs, with millions of people moving from place to place. Other pathogens such as Influenza A H1N1, in 2009, and various strains of the dengue virus, are known to have been introduced in the country through these hubs [7].

All countries that were strongly affected by COVID-19 have so far reported that the huge numbers of infected patients who require intensive care or who need to be admitted to a hospital have led the health care system into total collapse. Thus, knowing which regions in the country should be expected to be affected first, or harder, may help authorities to set priorities and take difficult decisions on how to better use scarce resources.

Here we analyzed the COVID-19 spreading risk within Brazil, taking the states of Rio de Janeiro and São Paulo as the starting points. In our analysis, we considered a scenario in which there are no mobility restrictions and as such configures the worst case scenario. Even though implementation of mobility restrictions and social distance are starting to be adopted by the country, it is still not clear whether the population will fully adhere to such restrictions, so that the current effect of these measures are not known yet.

Brazil presents strong spatial heterogeneity in terms of demography, age distribution, access to public health, and poverty indexes. Because of these inequalities, the COVID-19 epidemic should impact these populations differently, if factors such as transmissibility, lethality and vulnerability are taken into account.

In this article we provide quantitative metrics to help identify the regions in the country that are most vulnerable to the COVID-19. To identify these regions, we considered three main criteria: national mobility patterns, sociodemographic and economic characteristics of each region, and the health care infrastructure in terms of local hospitals capacity to handle the high demand that is expected for the COVID-19 pandemic.

## 2. Methods

### 2.1. Data

All analyses were done at the micro-regional administrative level. Brazil is divided into 558 micro-regional administrative units, whose sizes vary between 13 million people in the metropolitan area of São Paulo to 2, 703 in Fernando de Noronha island, in Pernambuco.

To measure the mobility between micro-regions, we used daily air travel statistics from the Official Airline Guide (OAG) [8] for long-range mobility, and shorter distance pendular travels from the 2010 national census (IBGE) [9].

For each micro-region, demographic data stratified by age were obtained from the 2000 and 2010 national censuses [9] and were used to project the population in 2020 per age group using a geometric growth model. From the Brazilian Institute of Geography and Statistics (*Instituto Brasileiro de Geografia e Estatística*, IBGE), the following socioeconomic indexes were collected: infant mortality, life expectancy, GINI index, human development index (education, longevity and income), proportion of individuals below the poverty threshold, proportion below the extreme poverty threshold, 25% percentile income, percentage of urban population, percentage of the population in households with piped water, % of population with insufficient water supply and precarious sewage disposal, and percentage of individuals in households without electricity. Infrastructure for COVID-19 hospitalization per micro-region was obtained 65 from DataSUS [10]. We calculated the number of standard hospital beds and complementary beds (Intensive Care Unit and Intermediate Unit) available for each micro-region [10], from the public (SUS) and private (non-SUS) sectors, per 10, 000 inhabitants.

### 2.2. Effective distance

To assess the probability of COVID-19 spreading within the Brazilian territory in the absence of mobility restrictions, we calculated the effective distance (*E*_*f*_ (*i, j*)) between micro-regions using air travel data. The value of *E*_*f*_ (*i, j*) is a measure of proximity between two micro-regions *i* and *j* created by the flow of people, which is known to strongly correlate with the time it takes to import infectious diseases into new territories from a well-defined origin [11, 12], particularly for diseases with direct transmission. To facilitate comparison between different scenarios, we also calculated a relative effective distance index (*e*_*f*_), dividing *E*_*f*_ by the distance to the nearest destination: *e*_*f*_ (*i, j*) = *E*_*f*_ (*i, j*)*/* min_*j*_*{E*_*f*_ (*i, j*)*}*. We computed *E*_*f*_ from São Paulo and Rio de Janeiro separately in order to assess the potential contribution of each one. This information was mapped into micro-region origin-destination pairs by summing over the corresponding airports serving each micro-region based on its municipality of reference.

### 2.3. Outbreak probability

To calculate the probability of outbreak in each micro-region *m*, we used the standard expression: 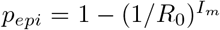 [13]. The prevalence of infection, *I*_*m*_, is estimated as *I*_*m*_ = *kτ*∑_*i*_ *f*_*i,m*_*I*_*i*_*/N*_*i*_, where *f*_*i,m*_ is the number of travelers with COVID-19 arriving from micro-region *i* to micro-region *m* while *I*_*m*_ is the product of the incidence *I*_*i*_*/N*_*i*_ times the infection duration *τ* and a scaling parameter *k*, to account for the number of undetected asymptomatic individuals participating in the transmission. The parameter *R*_0_ is the basic reproduction number [14]. For the purpose of the results shown, we set *R*_0_ = 2.5, which is compatible with previous studies [15, 16, 17, 18].

We computed the outbreak probability per micro-region using two scenarios:

#### First generation of outbreaks

We assumed community transmission taking place only in Rio de Janeiro and São Paulo, with 100 cases notified in a week at each municipality. Prevalence of infection was calculated by multiplying the notification count by an expansion factor *k* = 10 to take into account asymptomatic and unreported cases [19]. This number is then multiplied by the infection duration *τ* = 8 days [20], resulting in 8000 prevalent infections (infected *×* infection duration) in each of the two cities of origin.

The number of travelers per day between micro-regions was computed by adding the air travel data (used to calculate effective distance) and pendular mobility for work and study extracted from the 2010 Census [9]. The inclusion of pendular mobility is important to assess the spreading process between geographically close micro-regions, while still preserving data-driven flow estimates.

#### Second generation of outbreaks

In this scenario, we assumed that all micro-regions with *p*_*epi*_ *≥* 0.5 in the first scenario actually had outbreaks. In each of the outbreaks, the prevalence is set to the same level stated before for Rio de Janeiro and São Paulo micro-regions. Then we computed *p*_*epi*_ again, to identify the micro-regions most likely to develop outbreaks in this second round. These should not be confused with the infection serial interval.

It should be noted that this is a baseline scenario, which does not take into account the ongoing interventions affecting mobility, nor demographic and environmental effects that may play a role on the magnitude of *R*_0_. Our estimates describe the expected initial momentum of the spread in Brazil.

#### Social vulnerability

Social vulnerability is expected to affect the effectiveness of mitigation strategies to reduce morbidity and mortality. Partitioning cluster analysis (k-medoid) was performed to identify regions with similar social vulnerability. This method is a more robust variation of the standard k-means. The first step was the removal of highly linearly correlated variables. The following ones remained: life expectancy, GINI index, education component of the human development index, proportion of individuals below the extreme poverty threshold, percentage of urban population, percentage of the population in households with piped water, percentage of population with insufficient water supply and precarious sewage disposal, and percentage of individuals without electricity. The elbow method was used to determine the number of clusters. The analysis was done using the R environment [21], packages cluster [22], corrplot [23], FactoMineR [24], factoextra.

## 3. Results

### 3.1. Effective distance in the absence of travel restrictions

Figures 1(a-b) show the relative effective distance of Brazilian micro-regions from São Paulo and Rio de Janeiro generated by the typical travel movement by air. The majority of state capitals are among the closest areas, together with some important touristic destinations (such as Foz do Iguaçu/Paraná and Porto Seguro/Bahia), as well as important urban and industrial centers outside metropolitan areas such as Itajaí/Santa Catarina and Uberlândia/Minas Gerais. São Paulo shows a more central role than Rio de Janeiro, evidenced by the larger proportion of closer destinations, which means that it poses a greater risk for earlier and widespread case exportation to other states.

**Figure 1:**
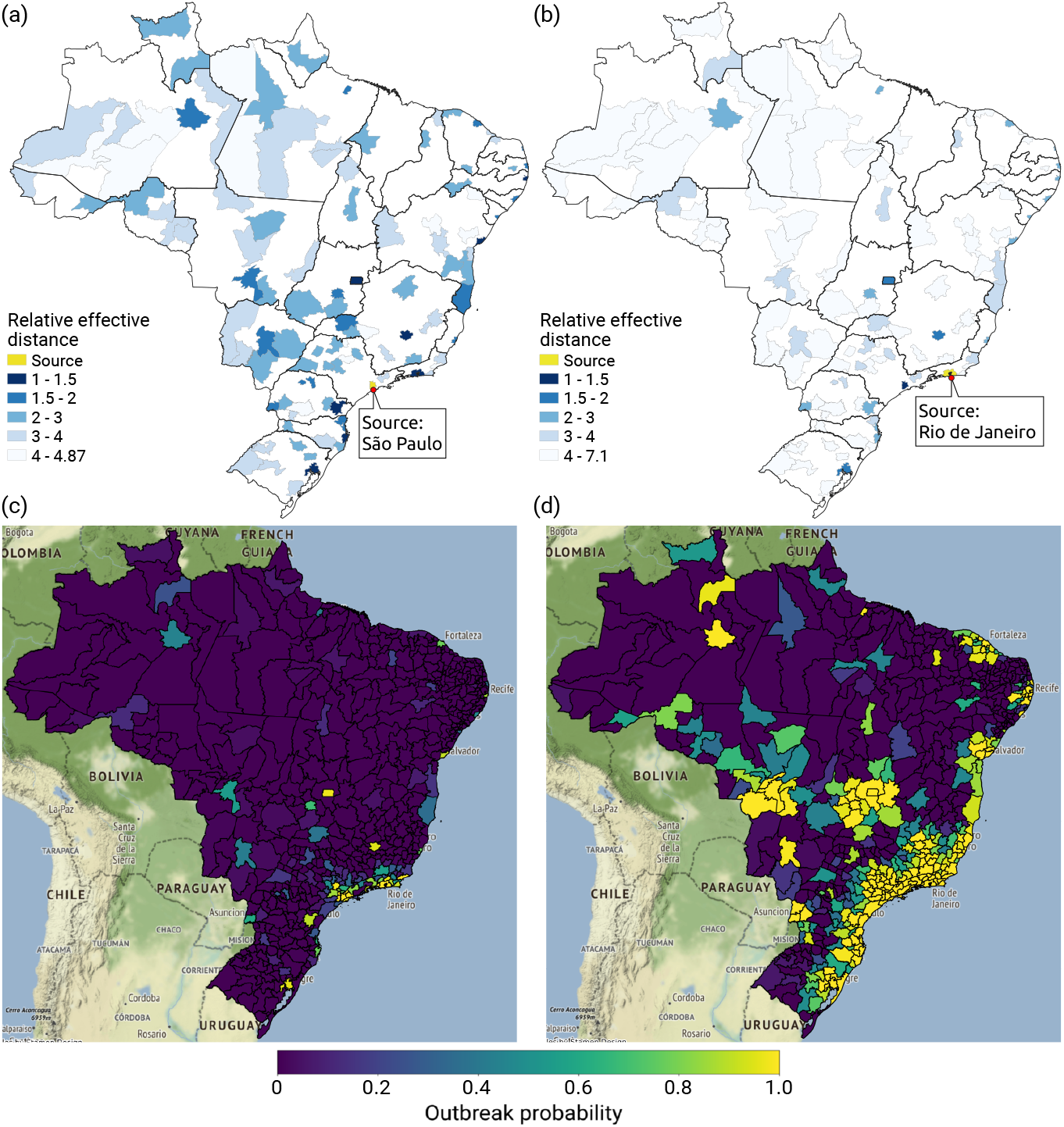
(a) Relative effective distance (*e*_*f*_) of Brazilian micro-regions from São Paulo based on airline network in the absence of travel restrictions; (b) the same from Rio de Janeiro, with blue gradient from closest (dark blue) to farthest (light blue) destinations, limited to those present on the airline network. Bottom panel uses both local and long-range mobility networks to estimate the (c) probability of COVID-19 outbreak per micro-region as Rio de Janeiro and São Paulo sustain high prevalence of infection; (d) second round of outbreaks after the micro-regions infected in (c) begin to contribute cases, with a gradient from dark purple (*p* = 0) to bright yellow (*p* = 1.0).

### 3.2. Probability of outbreak

Figure 1(c) shows the probability of outbreak triggered by the increased prevalence of COVID-19 in Rio de Janeiro and São Paulo, in the absence of travel restrictions. The most likely micro-regions to develop an outbreak are the geographic neighboring regions of São Paulo and Rio de Janeiro as well as all capitals in the South and Southeast regions (Belo Horizonte, Vitória, Curitiba, Florianópolis and Porto Alegre), Brasilia, Recife and Salvador, among others. A more complete list is found in the Supplementary material (Table S1).

Figure 1(d) shows the probability of outbreak in a secondary round of propagation, conditioned to the establishment of transmission in the micro-regions at highest risk (*p >* 0.5) during the first phase. In this second moment, the establishment of COVID-19 transmission is very likely in the majority of micro-regions along the coast, from Porto Alegre (in the South) to Salvador (North-east). Other high risk areas are the neighboring areas of Recife and Fortaleza (Northeast), the neighboring areas of Foz do Iguaçu, in the western border of Paraná state, and the neighboring areas of Cuiabá, Brasilia, and Goiânia in the Center-West.

### 3.3. Social vulnerability

We identified five classes of social vulnerability, tentatively ordered from A (least vulnerable) to D-E (most vulnerable) (1).

*Class A*. Mostly urban, with above-average life expectancy, comparatively less inequality, less population living in extreme poverty, better access to water supply and sewage disposal services, higher education. They are found in the largest cities and in the central region.

*Class B*. Very similar to A in life expectancy. Still more urban, but with more population living in extreme poverty (mean 5%). Inequality indexes and infrastructure are worse in comparison to A, but still above average. These are found in the South, Southeast, and central regions.

*Class C*. Mixture or urban and rural populations. In comparison to A and B, they have significantly lower life expectancy, significantly high poverty and less infrastructure. They are the most urbanized areas of the Northeast region. Manaus, capital of the Amazonas state in the North region, is also in this category.

*Class D*. Predominance of rural populations, high inequality, low HDIedu, poor access to water and sewage services, but with access to electricity. These are mainly located in the dry *Caatinga biome* area of the Northeast.

*Class E*. Predominantly rural regions in the Amazon. Low HDIedu, poor access to treated water, sewage disposal, and electricity.

Figure 2 shows four maps that synthesize the main vulnerabilities to COVID-19 in Brazil at this moment. Figure 2 (A) shows a strong difference in age structure, with the proportion of individuals with 60 or more years of age varying from only 3% in the North to more than 15% in the Southern part (Figure 2(A)). Figure 2 (C) shows micro-regions with similar social vulnerability indexes. Micro-regions in the C, D and E classes are the most vulnerable. They are located mostly in the Northeast and North regions. As expected, higher life expectancy is associated with better living conditions, significantly concentrated in the Southern portion of the country. Hospital capacity is very heterogeneous, the best capacity found in large metropolitan areas. There are unequipped micro-regions throughout the country but they concentrate in the North and Northeast.

**Figure 2:**
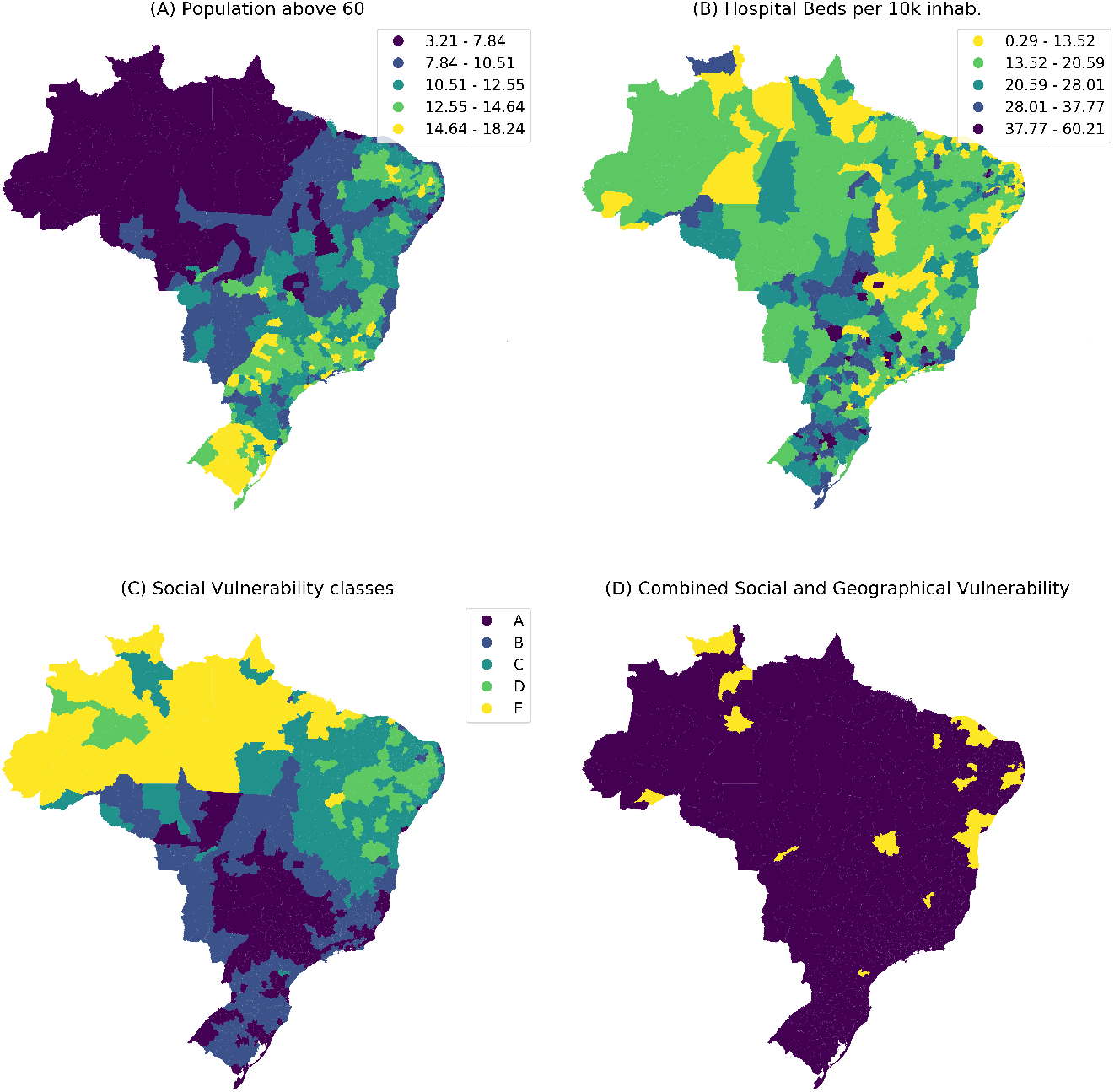
(Top left) Percentage of population above 60 years old (Top right) Hospital capacity as number of hospital beds per 10 per 10,000 individuals; (Bottom left) classification of homogeneous areas in terms of socio-economic vulnerability. D and E are the most vulnerable; (Bottom Right) selection of micro-regions with high probability of imminent COVID-19 epidemics and high social vulnerability.

Figure (D) 2 highlights the micro-regions with high probability of imminent outbreak (*p*_*out*_ *>* 0.5 on the second wave) and high social vulnerability. They are important targets for immediate attention (Supplement table S2) in terms of local socio-economic factors.

## 4. Discussion

In the absence of strong mobility restrictions, the fast establishment of COVID-19 outbreaks in the large urban areas of the country is highly likely, with subsequent spread to their vicinity municipalities. The time scale of this spread is not explicitly represented in the model, but it is a matter of a few weeks considering the short serial interval of this infection. The implementation of mobility restrictions can delay this spread but it is unlikely to change the routes of the travelling wave.

Social distancing is one of the main strategies put in place to delay transmission. This will be difficult to achieve in areas with high social vulnerability where poor living conditions will make it difficulty to adhere to hygiene and isolation protocols. Even in the *A* vulnerability class, the success of this strategy requires adaptation to cope with the large inequality within the municipalities of each micro-region. The classification proposed here can help to tailor the mitigation protocols to the needs and possibilities of the different regions.

It is clear the great heterogeneity in hospital capacity across the country. The median number of hospital beds is 19 per 10,000, but 5% of the micro-regions have only 6 beds per 10,000. This disparity poses an important challenge for resource allocation, and should be addressed as soon as possible specially for those municipalities combining high probability to early spread, relatively high percentage of individuals above 60 years old, and limited number of hospital beds per 10,000 people.

The regions in the North and Northeast of the country are those identified with the highest socio-economic vulnerabilities. These areas are expected to suffer above average burden if measures are not taken quickly, since the eventual disease spread would add to struggles already present in those populations.

Knowing in advance which regions can potentially suffer the biggest hit may allow authorities to opt for preemptive differential investments to the public health care system (SUS) in these regions. We hope the analysis presented here can help health authorities and decision-makers regarding the best course of action and how to better allocate their resources. Although analyses were carried on with Brazilian data alone, the approach presented here can be adapted for application in other countries as well.

## Data Availability

Data will become available after peer-review

## 5. Authors’ contributions

Mobility data analysis: MFCG, APP, JTD, AV

Outbreak probability analysis: FCC, CTC

Cluster analysis: OGC, RML

Study design, data interpretation, writing: FCC, RML, OGC, DV, LSB, CTC, MFCG

Revision of the final manuscript: all authors

## 6. Conflict of interest statements

We declare no conflict of interest

## 7. Role of funding source

The study was not funded by any source. Some of the authors have scholarships: D.V is a CNPq/Brazil fellow, RML is a Fiocruz fellow.

## 8. Ethics committee approval

Ethics committee approval not required. Data used in the study are aggregated without identification of individuals. They were obtained from official data sources.

## 9. Acknowledgments

We would like to thank Marcia Triunfol for her help in preparing this manuscript, Luiz Max Carvalho for his revision, and Alain Barrat for helpful discussions.

---

**Table 1:** Description of the five classes of social vulnerability. Life expect. = mean life expectancy (age), GINI = mean(GINI), poverty = % living in extreme poverty, water = % individuals without access to piped water, sewage = % population with insufficient water supply and precarious sewage disposal, electricity = % individuals in households without electricity, urban = % living in cities.

| Class | life<br>expect. | GINI | poverty | wa-<br>ter | sewage | electric-<br>ity | ur-<br>ban | HDI<br>edu. |
| --- | --- | --- | --- | --- | --- | --- | --- | --- |
| A | 75.0 | 0.46 | 2.16 | 95.9 | 1.30 | 0.39 | 0.79 | 0.64 |
| B | 74.2 | 0.49 | 5.99 | 89.15 | 4.10 | 1.59 | 0.64 | 0.57 |
| C | 70.9 | 0.52 | 20.38 | 80.46 | 15.266 | 4.27 | 0.56 | 0.50 |
| D | 69.96 | 0.53 | 25.72 | 61.88 | 24.14 | 4.03 | 0.49 | 0.47 |
| E | 70.82 | 0.60 | 31.55 | 70.09 | 41.15 | 16.84 | 0.50 | 0.45 |

## Notes

### Competing Interest Statement

The authors have declared no competing interest.

### Funding Statement

D.V. is a CNPq/Brazil Fellow, R.M.L is a INOVA/Fiocruz Fellow

